# Development and validation of a stakeholder-driven, self-contained electronic informed consent platform for trio-based genomic research studies

**DOI:** 10.1101/2024.05.01.24306461

**Authors:** Bethany Y Norton, James Liu, Sara A Lewis, Helen Magee, Tyler N Kruer, Rachael Dinh, Somayeh Bakhtiari, Sandra H. Nordlie, Sheetal Shetty, Jennifer Heim, Yumi Nishiyama, Jorge Arango, Darcy Johnson, Lee Seabrooke, Mitchell Shub, Robert Rosenberg, Michele Shusterman, Stephen Wisniewski, Blair Cooper, Erin Rothwell, Michael C Fahey, M. Wade Shrader, Nancy Lennon, Joyce Oleszek, Wendy Pierce, Hannah Fleming, Mohan Belthur, Jennifer Tinto, Garey Noritz, Laurie Glader, Kelsey Steffan, William Walker, Deborah Grenard, Bhooma Aravamuthan, Kristie Bjornson, Malin Joseph, Paul Gross, Michael C Kruer, the Cerebral Palsy Research Network.

## Abstract

Increasingly long and complex informed consents have yielded studies demonstrating comparatively low participant *comprehension* and *satisfaction* with traditional face-to-face approaches. In parallel, interest in electronic consents for clinical and research genomics has steadily increased, yet limited data are available for trio-based genomic discovery studies. We describe the design, development, implementation, and validation of an electronic iConsent application for trio-based genomic research deployed to support genomic studies of cerebral palsy. iConsent development incorporated stakeholder perspectives including researchers, patient advocates, institutional review board members, and genomic data-sharing considerations. The iConsent platform integrated principles derived from prior electronic consenting research and elements of multimedia learning theory. Participant comprehension was assessed in an interactive teachback format. The iConsent application achieved nine of ten proposed desiderata for effective patient-focused electronic consenting for genomic research.

Overall, participants demonstrated high *comprehension* and *retention* of key human subjects’ considerations. Enrollees reported high levels of *satisfaction* with the iConsent, and we found that participant *comprehension*, iConsent *clarity*, *privacy protections*, and *study goal explanations* were associated with overall *satisfaction*. Although opportunities exist to optimize iConsent, we show that such an approach is feasible, can satisfy multiple stakeholder requirements, and can realize high participant *satisfaction* and *comprehension* while increasing study reach.

## INTRODUCTION

Interest in the design and implementation of electronic informed consent platforms for both clinical care and biomedical research has been steadily increasing.^1^ Numerous challenges to obtaining truly informed genomic consents exist.^2^ Nevertheless, there is both growing interest in and utilization of electronic platforms for genomic research, although a number of important questions remain.

### Electronic consent use in clinical research and biobank studies

Over time, the need for full disclosure within traditional consents have led printed forms to become increasingly long and complex.^3^ When printed forms have been adopted to electronic formats, historically the long-form format is simply migrated to a tablet or similar device and reviewed by research coordinators during in person discussions. Research participants often do not fully comprehend information presented to them during the informed consent process.^4,5^ The updated Common Rule for human subjects research states that key information presented in a “concise and focused” manner will be most valuable to potential participants in deciding whether or not to join a clinical research study.^6^ Participant *satisfaction* with traditional long-form consent approaches is often relatively low but can be improved with use of simplified, straightforward language.^7^. In contrast, a biobanking study comparing electronic applications vs. long-form consents found that electronic consenting required not only significantly less staff time but was associated with improved participant *comprehension* of key elements of the informed consent.^8^

Electronic consents may also improve recruitment and retention in clinical research studies.^9^ This may be mediated in part by reducing access barriers for rural participants and by mitigating cultural and literacy barriers via the use of optional explanatory material.^10^ Assessments of stakeholders’ perspectives have also indicated that electronic frameworks can facilitate personalization and longitudinal interactions with research participants.^11^ Prior work has indicated that *satisfaction* is highly connected with ease of use and the overall participant experience. Mobile phone-supported electronic consents show a high degree of participant *satisfaction* and engagement,^12^ while a biobank electronic consent study demonstrated that testers overall had a positive experience with the portal but reacted negatively to an extensive identification verification process.^13^

### Electronic consent use in clinical genomic studies

Electronic informed consent has recently been expanded beyond biobanking studies to clinical genomic research by the Clinical Sequencing Evidence-Generating Research (CSER) consortium. The majority of participants found the approach acceptable and valued the access to testing that the study provided.^14^ CSER also assessed participants’ perspectives regarding electronic consenting for gene panel-based testing for eleven conditions estimated to affect 1-2% of the general population. The investigators found that participants’ decision to join the study was related to genetic self-efficacy, limited concerns about genetic screening, trust in the researchers, and the user-friendliness of the website.^15^

### Electronic consent use in genomic discovery research

Prior studies of electronic consent have been conducted in the context of the analysis of residual neonatal blood spots for broad research use, including genomic studies.^16^ This work has shown a high overall degree of participant *satisfaction* with electronic consent formats.^17^ Users have also highly rated the *clarity* and conciseness of information presented in electronic formats.^17^ Electronic consent utilization is also associated with higher participant *comprehension* of most key study concepts compared to long-form consents,^18^ particularly if teachback questions are employed to reinforce key study concepts.^19^

Based on the foundational evidence summarized above, electronic consents have now been designed and implemented for several major ongoing genomic research studies, including the Australian Genomics study,^20^ Simons Foundation’s SPARK study, the Autism Speaks MSSNG initiative, and the National Institutes of Health’s All of Us study.^21^ The All of Us study recently found that more than 95% of participants recognized the voluntary nature of the research study and were able to distinguish it from medical care.

### Hurdles to electronic consents

Although electronic consents for genomic discovery research are already being utilized, several important gaps in the field remain. In the context of discovery-based genomic studies (particularly those using trio-based designs), participant *satisfaction* and *comprehension* of key human subjects research elements have not been assessed.

Nevertheless, recognizing the potential inherent in electronic consents, Parra-Calderón and colleagues have proposed ten key elements (desiderata) necessary for the successful utilization of an electronic consent platform for genomic research (**Table 1**),^9,20,22–30^ although the challenges inherent in implementing these elements have not previously been overcome.

**Table 1.** Desiderata for Developing a Digital Genomic Informed Consent^a^.

| **Key Subject Areas and iConsent Goals** | **Integration of Desiderata in iConsent** |
| --- | --- |
| **Participant engagement:** Secure user interface enables continued and individualized consent management and engagement after enrollment <sup>20, 23, 24</sup> | • Interactive teachback questions<br>• Consent language downloadable<br>• Automated study status communications post-enrollment |
| **Personalized participation:** Transparency through provision of information at recruitment <sup>25, 26</sup> | • Clear explanation of risks, definitions, method of participant identification<br>• Opportunities to contact and engage with study team |
| **Participant information services:** Ability to monitor each researcher's access to data sets and biological samples over time <sup>27</sup> | • Impractical given widespread de-identified genomic data-sharing |
| **Healthcare integration:** For improved prevention, treatment, and care-delivery <sup>28, 29</sup> | • Optional return of results includes choice to share findings with participants and clinical providers |
| **Genomic literacy:** Facilitate understanding of precision medicine, genomic information, and implications of results <sup>28, 25</sup> | • Concise explanations within iConsent<br>• Teachback questions and 'More Info' tabs |
| **Incidental findings:** Bioethical principle of Autonomy supports individual's right to know and to not know <sup>25, 30, 27</sup> | • Incidental findings masked by bioinformatics workflow and not returned to participants; ethics committee engaged as appropriate to individual circumstances |
| **Digital accessibility:** iConsent accessible from all possible electronic formats <sup>9, 27</sup> | • Mobile, tablet, and laptop/desktop compatibility |
| **Identity management:** iConsent employs a trusted identity management process <sup>9, 27</sup> | • Secure user interface within REDCap incorporates authentication and authorization |
| **Audit trail:** Ability to reliably track consent status <sup>26</sup> | • Consent tracking status within REDCap framework |
| **Security and privacy:** Transparent protections <sup>9, 30</sup> | • Achieved via collaboration with IRB and implementation within iConsent framework |

<sup>a</sup>Desiderata derived from Parra-Calderón, et al.<sup>22</sup>

To address existing gaps, we sought to incorporate the desiderata while utilizing the concepts established by prior research. Accordingly, we describe the design, development, implementation and validation of a self-contained electronic iConsent application for trio-based genomic discovery research. This application is currently being utilized to enroll interested families in the Genetic Causes of Cerebral Palsy (GCCP) study conducted within the Cerebral Palsy Research Network (CPRN; www.cprn.org). We incorporated researcher, stakeholder, institutional review board, and genomic data-sharing perspectives and assessed *comprehension* and *satisfaction* among participants as well as potential factors driving these outcomes.

## PATIENTS AND METHODS

The GCCP iConsent was developed to support our ongoing study of cerebral palsy (CP) genomics. The study is being conducted at participating sites within the CP Research Network. Phoenix Children’s / University of Arizona College of Medicine – Phoenix serves as the genomics hub for this study. The study utilizes the CPRN’s clinical registry,^31^ which comprises >7,500 individuals with CP seen at centers across North America.

We designed our workflow so that enrolling in the GCCP study (overseen by the Phoenix Children’s central Institutional Review Board [IRB]) via the iConsent provides a secure means of obtaining informed consent. Participant-driven enrollment triggers sample collection via saliva kits mailed to the family’s home as a source of genomic DNA. After saliva samples are received for the trio, genomic sequencing (exome/ genome) and analysis is performed.

The CP Research Network utilizes dedicated clinical documentation templates integrated within the participating site’s electronic medical record. Clinical phenotypic data is entered by clinicians caring for patients in real time, and data is captured and securely transferred to the data coordinating center at the University of Pittsburgh.^32^ Participation in the CPRN clinical registry thus provides crucial phenotypic data. Privacy-preserving records linkage may then be used to link the clinical and genomic sequencing datasets, effectively connecting phenotype with genotype.

Since our study design is based on mother-father-child trios, in order to enroll the family, both biological parents are invited to participate given the high rate of *de novo* mutations in CP.^33^ This necessitated that the individual with CP, their biological mother, and their biological father each individually complete an assent/consent (as applicable). Under Arizona state law, legally authorized representatives are required to provide consent for those under the age of 18 and express permission is required for children 8-17 with the intellectual capacity to provide assent (as indicated by their legally authorized representative).

As we began the design of the iConsent application, needs assessments were conducted utilizing the Engage2020 Action Catalogue decision support tool.^34^ We adopted a community-based participatory action design framework and incorporated multiple stakeholder perspectives. Iterative consensus-building occurred using mixed methods approaches, including user committees and deliberative forums. Study information was provided to potential participants using an interactive webpage design (Wordpress) and study data were collected and maintained in a secure Research Electronic Data Capture (REDCap; Vanderbilt University)^35^ environment housed at the University of Arizona.

We used an embedded series of teachback questions to assess initial participant *comprehension* of core human subjects’ research concepts at the time of enrollment. We then deployed a follow-up survey to assess enrollees’ overall *retention* by re-presenting the teachback questions (**Supplemental Table 1**). We assessed *satisfaction* in the same survey, as well as factors that could influence *satisfaction*. Given that prior work has identified trust as a major factor influencing participation in CP genomic research studies,^36^ we assessed participant trust using the Hall Trust in Biomedical Research (H-TBR) scale, short form.^37^

**Statistical methods** *Comprehension* scores were calculated by summing the number of correct responses out of 5 and summarized by type of respondent (mother, father, or proband).

The distribution of initial and follow-up scores were compared by respondent type using two-tailed Wilcoxon rank sum tests. Responses to individual survey questions were tabulated by respondent type and compared using chi-square tests.

To evaluate pairwise differences in *comprehension* vs. *retention* (defined as continued understanding at the time of follow-up using the same questions initially presented) scores among the subset of individuals who completed both the initial and follow-up surveys, the comprehension score was stratified by respondent type and analyzed using Wilcoxon signed rank tests.

Kruskal-Wallis tests were performed to assess the relationship between participant *satisfaction* and other factors including *comprehension*, *retention*, *participant trust*, and measures of participants’ impressions of study *clarity*, *privacy protections*, explanations of *study goals*, and the *adequacy of information* provided regarding study participation. As the study was primarily designed to characterize responses by group, a formal sample size analysis was not appropriate. All GCCP study participants at least 18 years old and able to make independent medical decisions were invited to complete the initial survey. Of those, participants who returned a saliva sample were invited to complete the follow-up survey. All analyses were performed in SAS version 9.4 (SAS Institute, Inc).

## RESULTS

### I. Foundational principles, needs assessment, and design

#### Researcher perspective

A design committee was first assembled from research team members. This team identified key study goals and several important functions that the iConsent needed to fulfill. Fundamentally, the application needed to facilitate enrollment while preserving participant *comprehension* and maximizing *satisfaction* (here captured by the user experience). We also needed to generate metadata to connect members of the mother-father-child trio, coordinate sample collection, and generate unique identifiers to link de-identified participant phenomic and genomic data since these were being collected using different protocols at different sites. From a practical standpoint, our platform needed to be a) self-contained; b) professional in appearance (both inviting and secure); and c) user-friendly. We thus sought to balance competing needs – the need to generate a robust data set with the need to limit enrollee burden in order to facilitate participation. Instilling participant trust was also a primary goal of the study team.

As we conceptualized our iConsent, we sought to address existing gaps in the field.

Specifically, we sought to determine if a trio-based genomic research iConsent application that yielded high participant *comprehension* and *satisfaction* could be developed. We also set out to identify specific factors that influenced *comprehension* and *satisfaction* for the sake of subsequent quality improvement efforts.

With these principles in mind, we developed early study mock-ups. We scrutinized language and written communication heavily at these stages. We sought to explain the study opportunity, risks, and potential benefits to prospective participants using simplified language in an engaging, succinct format. Once the original study outline had been established, we then incorporated key elements related to community perspectives, human subjects’ protections, and data-sharing needs to further develop our study framework (**Supplementa**l **Figure 1**).

### II. Iterative development, incorporating stakeholder perspectives

#### Community perspective

We next incorporated perspectives from CP community and advocacy leaders by partnering with the community advisory committee of the CPRN. This stakeholder group included individuals with CP as well as parents of a child with CP to represent potential participants. We solicited feedback through a brief survey, inviting impressions of our early mock-ups. Comments and suggestions varied among group members, but several important themes emerged to optimize the user experience and thus overall *satisfaction* (**Table 2**). Community feedback prompted us to improve the user interface to create a more intuitive and easily navigable iConsent. Recommendations for clear and concise language were also incorporated into evolving study mock-ups before presentation to our institutional review board.

**Table 2.** Incorporated Recommendations from Community Stakeholders, IRB members, and Mayer’s Multimedia Learning Theory.

| Topic | Incorporated elements |
| --- | --- |
| **Community stakeholder recommendations** |  |
| Language | Ensure language is concise, consistent, and understandable to target audience |
| User experience | Revise graphical user interface to be more intuitive |
| Stress key study elements | Emphasize key aspects of the study via placement and presentation |
| Clarify trio enrollment study design | Emphasize that all three family members need to individually enroll in order for the family unit to participate (given that key information often needs to be presented multiple times in different ways) |
| Assent as appropriate | Explain to parent why their child needs to provide assent |
| **IRB recommendations** |  |
| Communication of study information | Balance brevity with full disclosure and detailed study information through the employment of 'More Info' tabs within the application<br>Optimize language to ensure it is at a 6th grade reading level |
| Risk/benefit consideration | Convey what participation entails, ensuring potential benefits and/or risks of participation are outlined before informed consent is obtained |
| Comprehension | Ensure teachback questions assess understanding of key aspects of participation |
| Documentation of informed consent | Provide copies of electronically signed, time stamped informed consents to enrolled participants |
| **Principles of Mayer's Multimedia Learning Theory<sup>a</sup>** |  |
| Coherence | Concise language was used |
| Signaling | Cues oriented users to essential material (bold typeface, 1-2-3, etc.) |
| Redundancy | Reiteration was limited to emphasizing key concepts |
| Spatial contiguity | Text was grouped for user-friendliness; bullet points employed |
| Temporal contiguity | Related text and images were presented together |
| Segmenting | User-paced pages partitioned material; ability to pause and return |
| Pretraining | Introductory video provided study overview |
| Modality | Graphics and narration were utilized |
| Multimedia | Text, graphics, video, and human voice were integrated |
| Personalization | Conversational style employed throughout |
| Voice | Human voice-over used in video narration |
| Image | Unnecessary images were avoided to focus on key content |
| Convergence between community stakeholder and IRB recommendations with concepts of Mayer's Multimedia Learning Theory was noted, especially relating to language use and presentation of information. Early IRB and stakeholder engagement yielded valuable suggestions with actionable solutions, while integration of Mayer's principles further advanced communication targets prioritized in community feedback. |  |

<sup>a</sup> Principles derived from Mayer's Cognitive Theory of Multimedia Learning<sup>42</sup>

### IRB perspective

Electronic consents had not previously been utilized at our institution, so we partnered with our IRB to ensure that robust human subjects protections were incorporated into iConsent. We relied on established principles that provision of informed consent requires decision-making capacity, voluntariness, *adequate information*, and *comprehension*.^2^ We presented our mock-ups via a series of consensus-building deliberative forums with IRB leaders that led to further refinements. IRB leaders emphasized that key informed consent concepts needed to be presented in sufficient detail to be understood by those signing the informed consent. They were enthusiastic about the use of embedded teachback questions to ensure *comprehension* and *retention* of these key study concepts in order for the iConsent to represent a valid alternative to traditional in-person, long-form based informed consent. Important concepts derived from these discussions are outlined in **Table 2**.

### Data-sharing considerations

Worldwide, data-sharing concepts have become an important part of genomic research, as combining cohorts can increase statistical power and facilitate discovery. In the United States, data-sharing has been a priority topic for the National Institutes of Health (NIH) and has become an integral part of federally-funded genomic research. We sought to ensure that our study was aligned with the latest recommendations from the NIH.^38^ We incorporated current guidelines for precise informed consent^6^ in order to maintain transparency and facilitate *trust* between researchers and study participants while still enabling

high-level data-sharing for genomic discovery.^39^ This resulted in additional meetings with IRB leaders and several additional rounds of revision, including the incorporation of broad data and sample sharing for research use as an intrinsic aspect of study participation. Informed consent language was expanded to detail data and sample sharing, informational content was added, and the interactive teachback questions were modified to promote participant *comprehension*.

### III. iConsent implementation

Our iConsent platform used a rich media format developed in a WordPress environment that presented information about the study and requirements for participation (**Supplementa**l **Figure 2**). We incorporated multimedia elements given evidence that this improves participant experiences.^40^ We used a combination of stock photos and photographs submitted to both the LifeShots competition (sponsored by the American Academy of Cerebral Palsy & Developmental Medicine) and CPRN Photo Contest after obtaining appropriate permissions.

We produced a short animation (Video Animation, Inc) providing an overview of the study that was less than 2 minutes in length but encapsulated crucial aspects of participation (**Supplementary Video**). The second portion of the application utilized RedCap to securely collect participant personally identifying information.

#### Content development

We sought to satisfy human subjects regulations while balancing the needs of our study population and research goals. We incorporated elements of cognitive load theory^41^ to lessen participant burden and reduced individual options to customize participation as compared to our original long form consent. We then incorporated elements of Mayer’s multimedia learning theory (**Table 2**),^42^ which has been reported to improve both *comprehension* and *satisfaction*^43^ in the development of the iConsent. We anticipated that participant preferences for information about aspects of study participation would vary, but anticipated that presenting too much information in too short a time frame would reduce *comprehension*. Therefore, we identified crucial human subjects research elements that represented core participant-facing material (see **Table 2**) and then provided “More Info” tabs for those interested in obtaining more detail. Additional community perspectives were valuable at this point as we sought to strike a balance between thoroughness and brevity to yield clear, concise language.^44^

### Final review and integration

After incorporating these changes, the research team internally re-reviewed the iConsent. We then confirmed the suitability of the application’s content for launch via a final review with IRB leaders. Both of these measures were taken to ensure that all of the critical concepts we identified during the course of study development were faithfully incorporated and all concerns were addressed and recommendations incorporated to the best of our ability. In cases where recommendations were at odds (i.e. remove a specific element vs. retain that element) final decisions were made by the study team in a consensus-building fashion.

After extensive internal beta testing, we launched the iConsent in March 2021. Potential participants were invited to participate in the study by email, study brochure, study poster, or direct mailing if they received care at a participating center (AI duPont Hospital for Children, Colorado Children’s, Nationwide Children’s, Phoenix Children’s, Seattle Children’s, St. Louis Children’s, or the University of Texas – Houston). We tracked iConsent completion rate (full trio enrollments) using an internal tracking system and developed an automated text messaging tool to help reduce the proportion of incomplete trios. We adopted our iConsent to iOS, receiving approval to deploy the app from Apple, Inc. and launched iConsent for iPad August 2022.

### IV. iConsent validation

#### User experience and implementation

Of the 561 families contacted, 458 (81.6%) returned at least one survey from a family member. Data from 821 participants was available to assess comprehension (380 fathers, 420 mothers, and 21 probands). Responses to teachback questions indicated good overall *comprehension* of key human subjects’ considerations, with a mean score = 4.1 ± 1.1 (out of 5), including the voluntary nature of participation, potential risks of participation, study goals, and privacy considerations (**Figure 1**, **Supplemental Figure 3)**.

**Figure 1.**
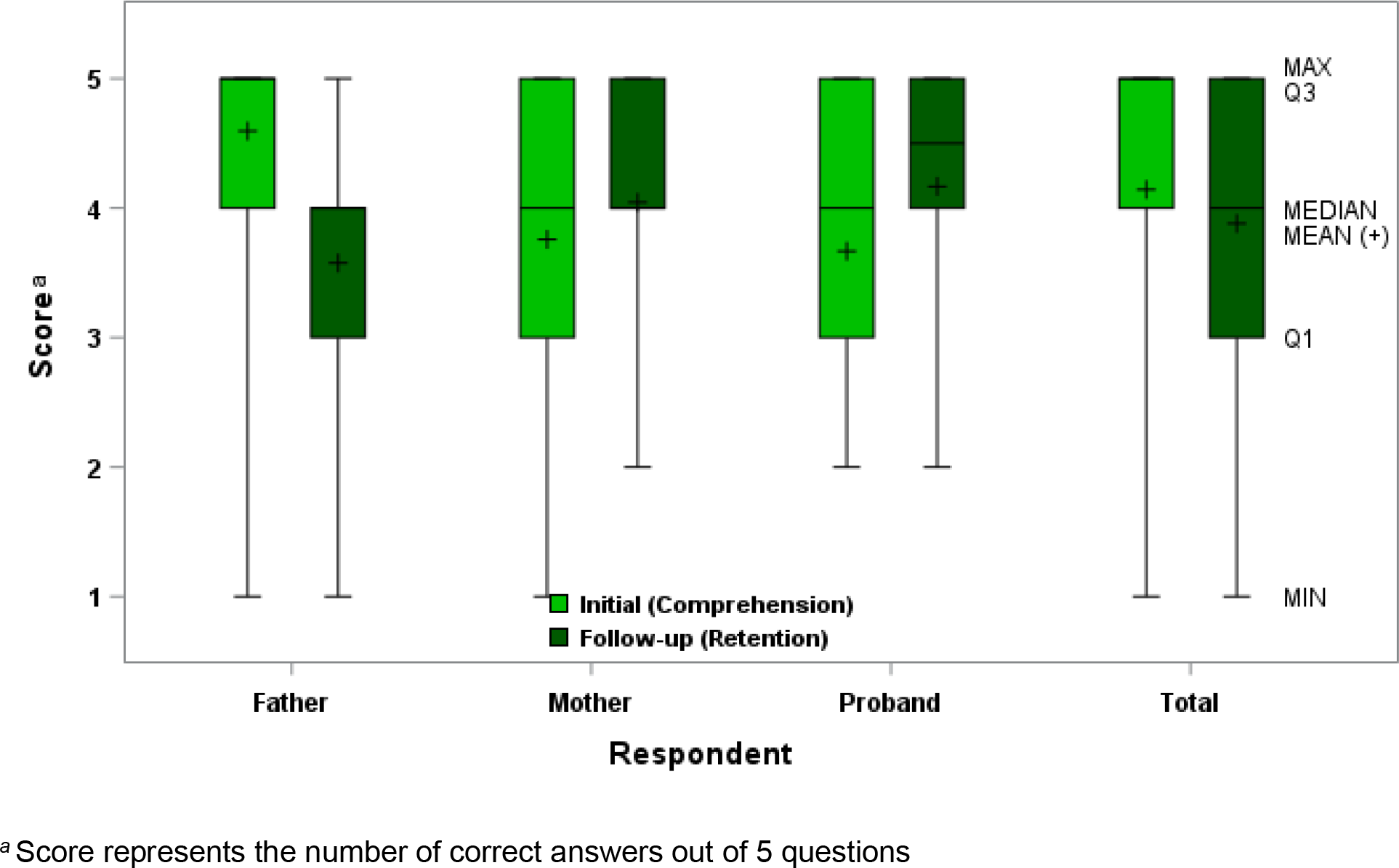
Overall Comparison of Initial (C*omprehension*) and Follow-up (R*etention*) Scores

However, we noted that many participants, particularly mothers, answered Question 1 “Do I have to participate in the genetic causes of CP study?” incorrectly (**Supplemental Table 2**). We solicited qualitative feedback from those that answered incorrectly, allowing us to identify a common theme – many parents interpreted this question as an inquiry as to whether their participation was needed to complete the trio. Consequently, we rephrased Question 1 to read “Is it my choice to be part of the Genetic Causes of CP Study?” This led to a higher correct response rate (99.2% for the rephrased question vs. 56.8% for the initial question; p <0.0001).

#### Follow-up Survey

After participating families enrolled in the study and provided saliva samples, they received a follow-up survey. Nearly forty-two percent of enrolled families (142/340) responded to the follow-up survey. The majority of respondents (>90% in each instance) indicated that they were *satisfied* with their experience (assessed by ease of use of the iConsent), that they were given *adequate information* about the study to make an informed decision to participate, that the *study goals* were clear, that the iConsent was *clear*, and that *privacy protections* were sufficient (**Figure 2**).

**Figure 2.**
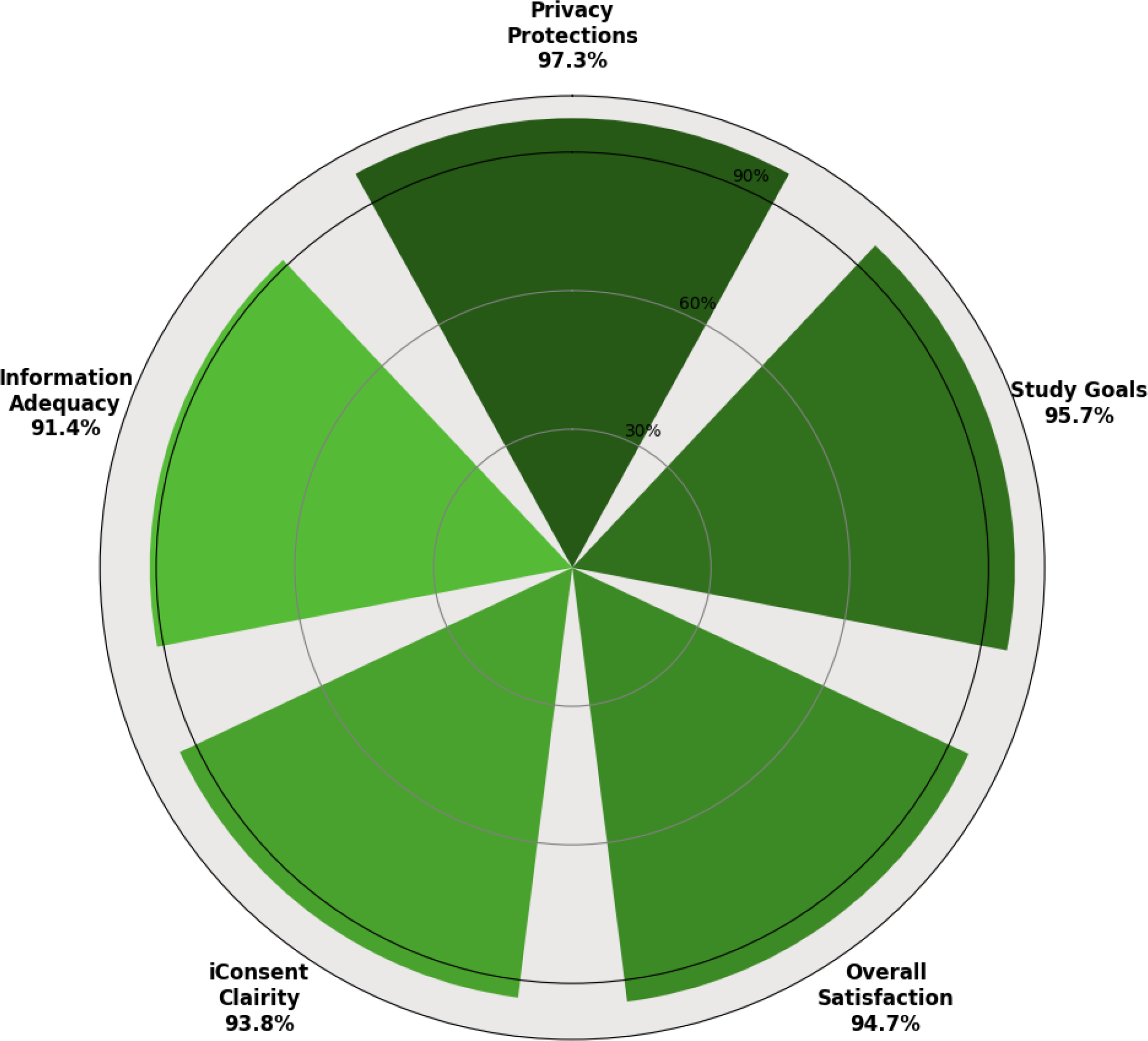
Overall Participant *Satisfaction* and Responses for Key Elements Impacting *Satisfaction*. Darker shades represent higher degrees of *Satisfaction*

We assessed participant *retention* by comparing initial teachback scores to follow-up survey teachback scores (the same questions were asked in both instances). We found a small but statistically significant dropoff in *retention* (mean initial *comprehension* = 4.1 ± 1.1 vs. mean *retention* = 3.9 ± 1.0; p<0.0001 by Wilcoxon signed rank test). However, *retention* remained relatively high even after a mean of 101.7 days ± 116.1 had elapsed since initial enrollment. We assessed the potential impact of the modification we made to Question 1 via a sensitivity analysis, which demonstrated that this change did not substantially impact our overall interpretations (**Supplementary Methods and Results**).

Finally, we then turned our attention to the factors that may have influenced overall *satisfaction*. We found that study *comprehension*, *iConsent clarity*, *privacy protections*, and appropriate explanations of *study goals* were associated with participant *satisfaction* (**Figure 3**, **Supplemental Table 3**). The *adequacy of information* presented was equivocally associated with *satisfaction* (p = 0.0552). Conversely, no relationship was found between *retention* of key concepts and overall *satisfaction*. Furthermore, no association between *participant trust* and *satisfaction* was evident.

**Figure 3.**
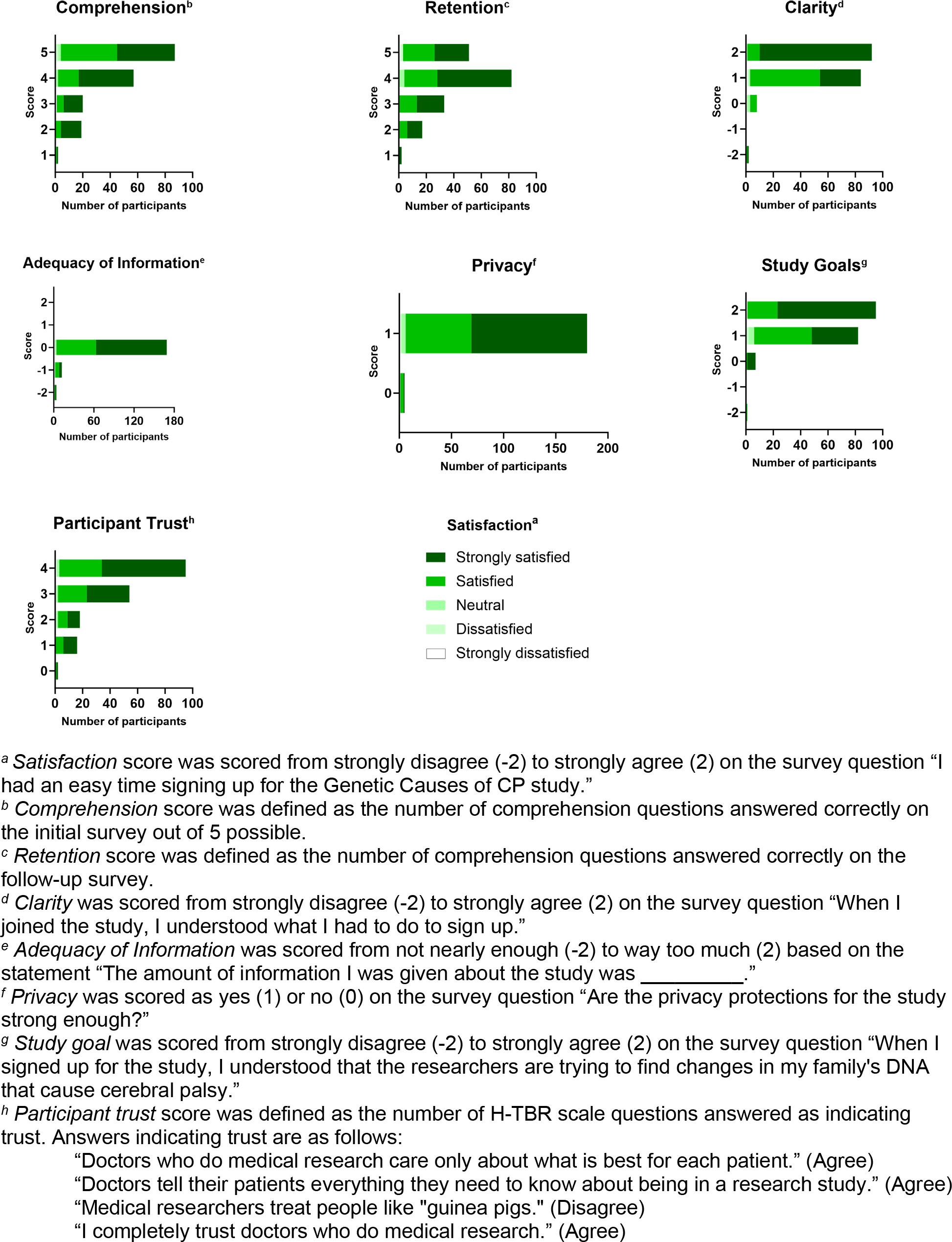
Association Between Study Elements and Overall *Satisfaction* Scores

## DISCUSSION

We describe the design, development, implementation, and validation of a fully self-contained electronic iConsent application suitable for multi-institutional recruitment of mother-father-child trios to participate in genomic discovery-based research. Although our study was targeted to families with a child with CP, the iConsent was designed to be modular enough to readily adapt to other applications. We incorporated elements identified as important by broad stakeholder representation, including the research team, CP community, IRB, and federal agencies. Our findings of high participant *comprehension* and *retention* as assessed by teachback questions served as a key validation of our approach. We also implemented nine out of the ten desiderata that had previously been proposed for electronic genomic research consents (**Table 1**). We did not incorporate the participant information management consideration given that we deemed it impractical to notify participants of every occasion when their data were utilized, particularly if their data became part of a large-scale de-identified genomic database as is typical for current genomics research.

We developed and validated our application as a standalone tool to increase access to research while preserving participant *comprehension* and *satisfaction*. More than 95% of participants indicated that they felt the iConsent was *clear*, and >90% of participants also reported that it contained an *adequate amount of information* for them to make an informed decision. These findings suggest that the multimedia structure we adopted and the learning principles that we utilized were effective in delivering digestible content, as indicated by the overall high participant *comprehension* scores.

Given the design of our study, we were not able to formally compare our outcomes to traditional paper-based in-person consenting, representing a potential limitation of our findings. Accessibility may be somewhat lesser for those of lower socioeconomic status as our application requires a device with internet connection and is currently only available in English, although a Spanish language format is in development. As SES, education level, and literacy were not the foci of this study, data on these metrics was not collected. Nevertheless, as prior studies have indicated, participant decision aids may be useful in addressing potential barriers to participation in the future.^45^

Our prior work with stakeholders (families with CP) suggested that the most important factors that might influence their willingness to participate in genomic research included (i) a clear understanding of *study goals*; (ii) adequacy of *privacy protections*; and (iii) *trust* in biomedical researchers.^36^ Here, when we surveyed participants who had already enrolled in CP genomic research, we confirmed that understanding *study goals* and sufficient *privacy protections* were both associated with overall *satisfaction*, while *participant trust* was not identified as a factor influencing *satisfaction*. This may reflect subtle differences in families’ trust of biomedical researchers in general vs. those focused on salient topics (in this case, CP). It is possible that a certain threshold of *trust* is needed for participants to enroll. These findings could also indicate that *trust* does not influence behavior as much as it may reflect existing attitudes.

We found that the participant *comprehension*, iConsent *clarity*, *privacy protections*, and *study goal explanations* all contributed to overall *satisfaction*. Our findings indicate that these elements should be considered to design effective electronic consents for genomic research.

Future research will include opportunities to optimize the presentation of human subjects’ research concepts and further improve *comprehension* and *retention*. Additional updates we plan to implement are anticipated to further enhance the user experience via a personalized study dashboard and facilitate ongoing communication with the study team using text messaging and automated updates to further align researchers and participants for mutually beneficial clinical research studies.

## Data Availability

Participant informed consent authorizes sharing of de-identified data with researchers for research purposes only. Anonymized survey data will be made available upon reasonable request to the corresponding author.

## Supporting information

Supplementary Video - Brief Animated Study Overview

## Data Availability

https://www.nodataavailabilityurl.com

## Acknowledgments

We are grateful to the families within the cerebral palsy community who have supported this work, particularly members of the CP Research Network community advisory council who provided critical feedback in the initial stages of this work. We appreciate the contributions of Jacob Kean to initial discussions of the iConsent and the input of Steve Esquivel regarding design considerations for the iConsent. We particularly wish to thank Bill Lewis, who provided crucial logistical and technical feedback and support for this project from conceptualization through implementation.

## Author Contributions

1. Conceptualization: M.C.K., J.L., S.A.L., H.M., T.N.K., R.D., S.B., S.N., S.S., J.H., Y.N., J.A., D.J., L.S., M.S., R.R., M.S., S.W., B.C., E.R., M.F., CPRN Investigators, M.W.S., N.L., J.O., W.P., H.F., M.B., J.T., G.N., L.G., K.S., W.W., D.G., B.A., K.B., P.G., B.Y.N.
2. Data curation: M.C.K., J.L., S.A.L., H.M., T.N.K., R.D., S.B., S.N., S.S., B.Y.N.
3. Formal analysis: M.C.K., S.S., M.J.
4. Funding acquisition: M.C.K.
5. Investigation: M.C.K., J.L., S.A.L., H.M., T.N.K., R.D., S.B., S.N., S.S., B.Y.N.
6. Methodology: M.C.K., J.L., S.A.L., H.M., T.N.K., R.D., S.B., S.N., S.S., Y.N., J.A., D.J., L.S., M.S., R.R., M.S., B.C., E.R., M.F., CPRN investigators, B.Y.N.
7. Project administration: M.C.K., J.L., H.M., B.Y.N.
8. Resources: M.C.K.
9. Software: M.C.K., J.L., H.M., T.K., B.Y.N.
10. Supervision: M.C.K., J.L., B.Y.N.
11. Validation: M.C.K., J.L., H.M., T.K.
12. Visualization: M.C.K., J.L., H.M., T.K., S.S., S.B., B.Y.N.
13. Writing-review & editing: M.C.K., J.L., S.A.L., H.M., T.N.K., R.D., S.B., S.N., S.S., J.H., Y.N., J.A., D.J., L.S., M.S., R.R., M.S., S.W., B.C., E.R., M.F., CPRN Investigators, M.W.S., N.L., J.O., W.P., H.F., M.B., J.T., G.N., L.G., K.S., W.W., D.G., B.A., K.B., P.G., B.Y.N.

## Ethics Declaration

Phoenix Children’s IRB reviewed and approved this study as the single IRB (sIRB). Informed consent was obtained from all participants. Although participant level phenotypic data were not included in our analysis, these de-identified data were obtained for the trio-based genomic study through collaboration with the CPRN registry.

## Supplementary Materials

**Supplemental Table 1.** Follow-up Survey Questions

**Supplemental Figure 1.** Stakeholder Perspectives. Integrated needs analysis incorporating researchers, IRB, community, and data-sharing perspectives.

**Supplemental Figure 2.** Electronic iConsent App Representative Image

**Supplementary Video.** Brief Animated Study Overview

**Supplemental Figure 3.** Distribution of Initial (Comprehension) and Follow-up (Retention) Scores

**Supplemental Table 2.** Overall Initial and Follow-up Question Responses – Breakdown

**Supplementary Methods and Results.** Sensitivity Analysis and Comparison of Initial and Follow-up Scores to Survey Question 1

**Supplemental Table 3.** Association Between Study Elements and Overall Satisfaction Scores (Expanded Data)

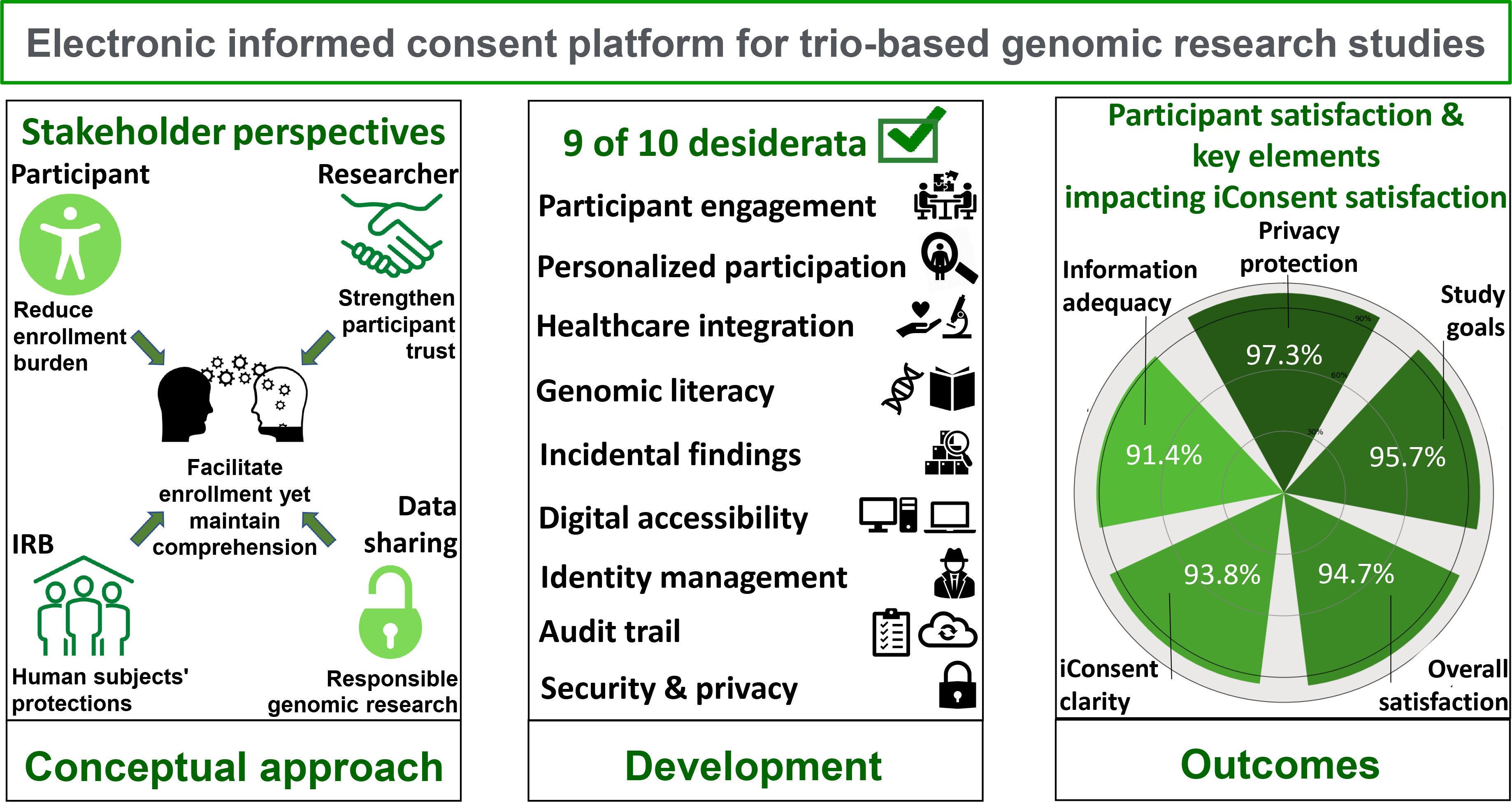

