## Supplementary Materials - Tables and Figures for "Development and validation of a stakeholder-driven, self-contained electronic informed consent platform for trio-based genomic research studies"

**Supplemental Table 1.** Follow-up Survey Questions

| Metric | Survey question |
|---|---|
| **Overall satisfaction** | 1. I had an easy time signing up for the Genetic Causes of CP study (strongly agree, agree, uncertain, disagree, strongly disagree). |
| **Comprehension (initial)**<br>**retention (follow-up)**<br><br>(sum #2-6) | 2. Do I have to participate in the Genetic Causes of CP study? (Yes, No)<br>3. Are there any risks involved in participating in the study? (Yes, No)<br>4. Will the information learned in this study go on to help people in the CP community? (Yes, No)<br>5. My samples and data may be shared with anyone who wants to use them, for any reason. (Yes, No)<br>6. Other researchers who use my samples and data in the future will be provided with my personal information like my name, phone number, and address. (Yes, No) |
| **Adequacy of information** (#7)<br><br>**Study clarity** (#10)<br><br>**Privacy** (#11) | 7. The amount of information I was given about the study was _________ (way too much, too much, just enough, not enough, not nearly enough)<br>8. How can we make the study better?<br>9. How can we make it easier to sign up for the study?<br>10. When I joined the study, I understood what I had to do to sign up (strongly agree, agree, uncertain, disagree, strongly disagree)<br>11. Are the privacy protections for the study strong enough? (Yes, No)<br>11-a. If not, what would help you feel more comfortable? |
| **Study goals** (#12) | 12. When I signed up for the study, I understood that the researchers are trying to find changes in my family's DNA that cause cerebral palsy (strongly agree, agree, uncertain, disagree, strongly disagree). |
| **Participant trust**<br><br>(sum #13-16) | 13. Doctors who do medical research care only about what is best for each patient (agree, disagree).<br>14. Doctors tell their patients everything they need to know about being in a research study (agree, disagree)<br>15. Medical researchers treat people like "guinea pigs."<br>16. I completely trust doctors who do medical research (agree, disagree) |

**[a]Too much indicates not concise; too little indicates not thorough**

**Supplemental Figure 1.** Stakeholder Perspectives

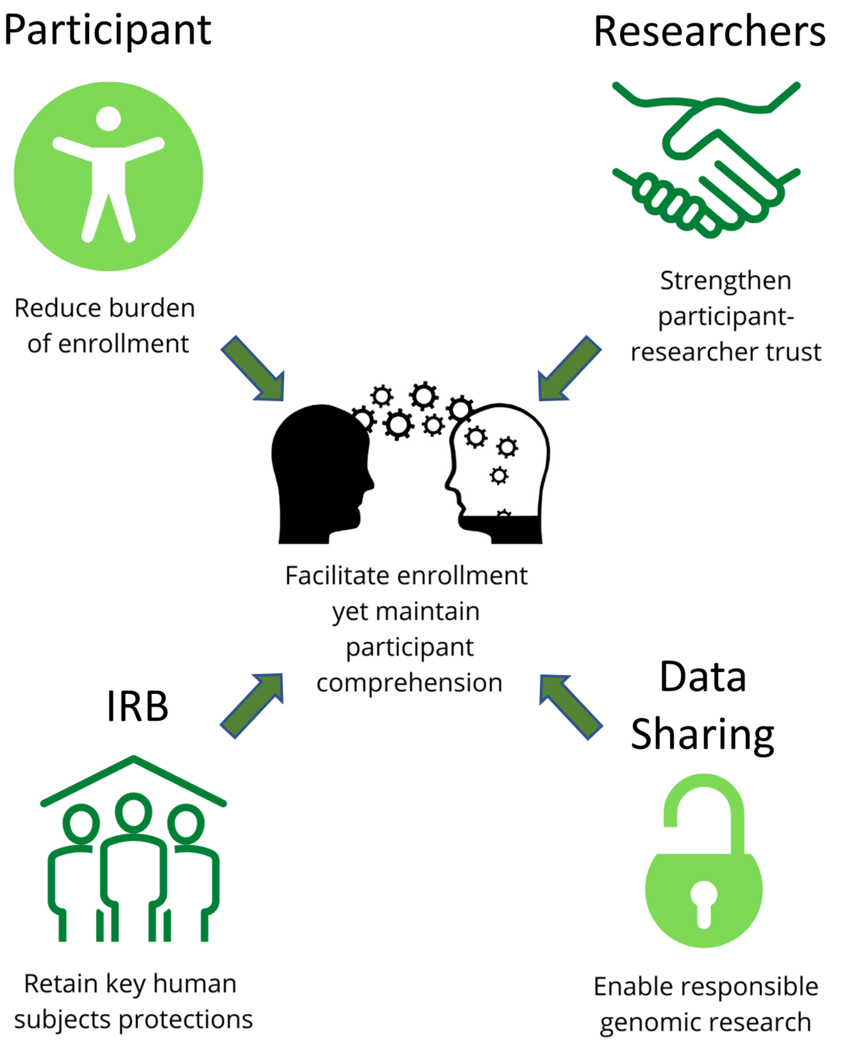

**Supplemental Figure 2.** Electronic iConsent App Representative Image

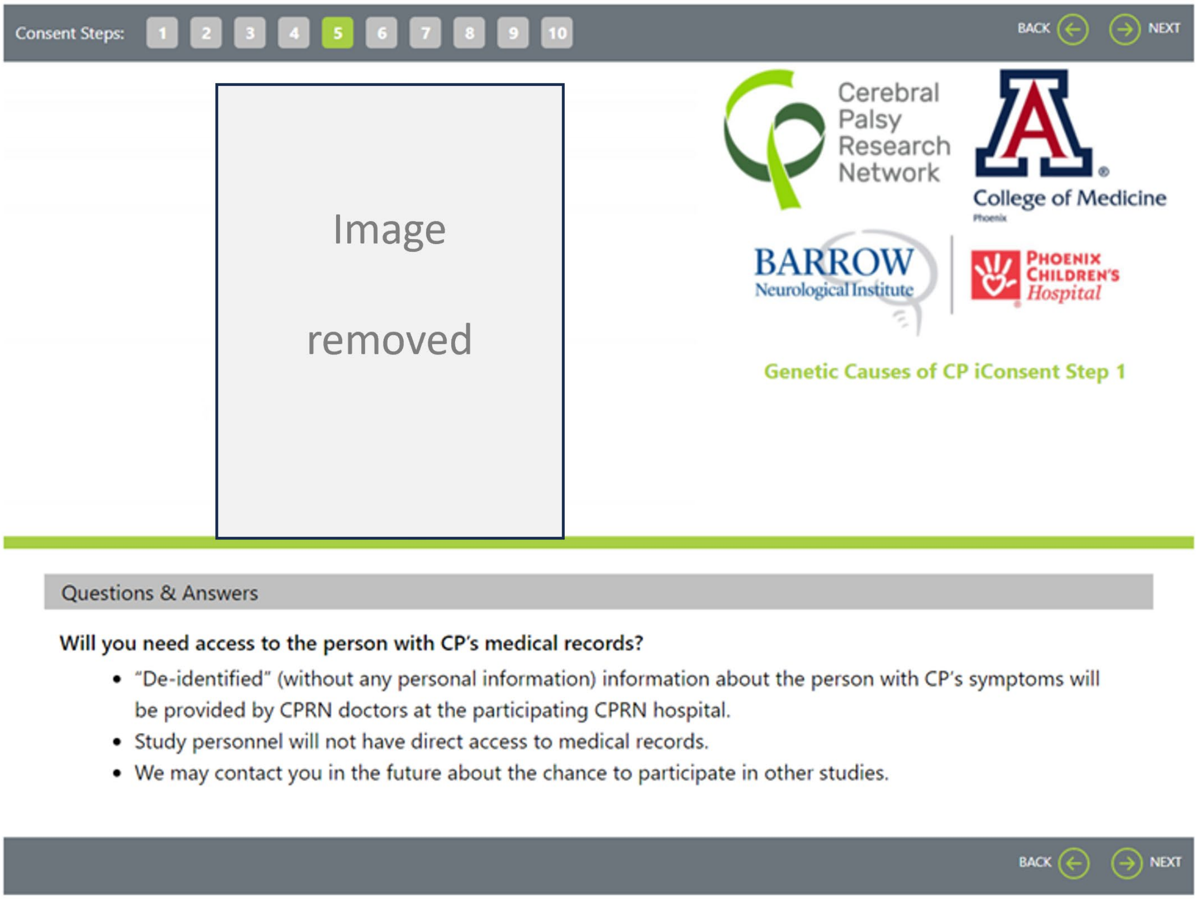

**Supplemental Figure 3.** Distribution of Initial (Comprehension) and Follow-up (Retention) Scores[a]

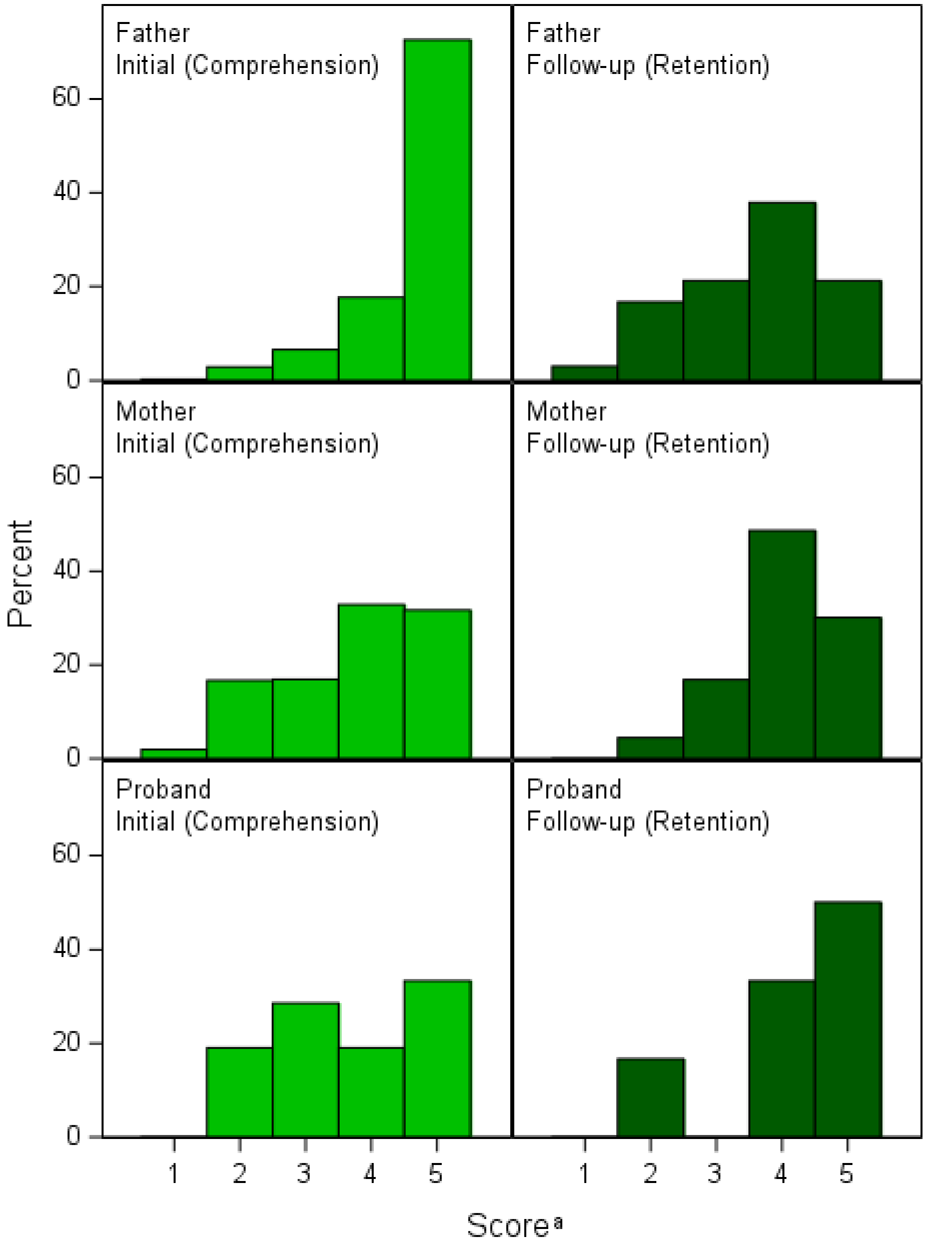


[a] Score represents the number of correct answers out of 5 questions

**Supplemental Table 2:** Overall Initial and Follow-up Question Responses – Breakdown

| | | Fathers | | | Mothers | | | Probands | | | Total | | |
|---|---|---|---|---|---|---|---|---|---|---|---|---|---|
| **Survey question** | **Correct response** | **Initial N=380** | **Follow-up N=66** | **pvalue[a]** | **Initial N=420** | **Follow-up N=113** | **pvalue[a]** | **Initial N=21** | **Follow-up N=6** | **pvalue[a]** | **Initial N=821** | **Follow-up N=185** | **pvalue[a]** |
| Do I have to be part of the GCCP study?[b] | No | N=164<br>148 (90.2%) | N=29<br>23 (79.3%) | 0.0877 | N=183<br>104 (56.8%) | N=53<br>48 (90.6%) | <0.0001 | N=10<br>10 (100.0%) | N=4<br>4 (100.0%) | . | N=357<br>262 (73.4%) | N=86<br>75 (87.2%) | 0.0070 |
| Is it my choice to be part of the GCCP study?[b] | Yes | N=216<br>212 (98.1%) | N=37<br>37 (100.0%) | 0.4041 | N=237<br>235 (99.2%) | N=60<br>57 (95.0%) | 0.0254 | N=11<br>11 (100.0%) | N=2<br>2 (100.0%) | . | N=464<br>458 (98.7%) | N=99<br>96 (97.0%) | 0.2109 |
| Are there risks involved in participating? | Yes | 330 (86.8%) | 14 (21.2%) | <0.0001 | 224 (53.3%) | 39 (34.5%) | 0.0004 | 12 (57.1%) | 3 (50.0%) | 0.7562 | 566 (68.9%) | 56 (30.3%) | <0.0001 |
| Will the information learned in this study be used to help people in the CP community? | Yes | 367 (96.6%) | 65 (98.5%) | 0.4124 | 416 (99.0%) | 112 (99.1%) | 0.9474 | 19 (90.5%) | 6 (100.0%) | 0.4321 | 802 (97.7%) | 183 (98.9%) | 0.2892 |
| My samples and data may be shared with anyone who wants to use them, and for any purpose. | No | 350 (92.1%) | 49 (74.2%) | <0.0001 | 303 (72.1%) | 106 (93.8%) | <0.0001 | 12 (57.1%) | 5 (83.3%) | 0.2414 | 665 (81.0%) | 160 (86.5%) | 0.0792 |
| Other researchers who use my samples and data in the future will be provided with my personal information like my name, phone number, and address. | No | 339 (89.2%) | 48 (72.7%) | 0.0003 | 296 (70.5%) | 95 (84.1%) | 0.0037 | 13 (61.9%) | 5 (83.3%) | 0.3261 | 648 (78.9%) | 148 (80.0%) | 0.7459 |

[a] Chi-square test
[b] Question was changed during the study; participants only answered one version of the question

**Supplementary Methods and Results.** Sensitivity Analysis and Comparison of Initial and Follow-up Scores to Survey Question 1

**Sensitivity analysis** Given that many respondents were answering question 1 (Q1) incorrectly, the question was revised after qualitative analysis. This resulted in 34 respondents receiving one version for their initial survey and a different version for their follow-up survey. A sensitivity analysis was thus conducted to see whether the results were similar when these respondents were removed.

In the sensitivity analysis, 324 people were given the original version of the question on their initial surveys and 463 were given the updated version. On follow-up, 85 got the original and 66 got the updated version. All groups improved at follow-up except for fathers who had the original version of the question; 89.5% of them answered correctly on the initial survey and 79.3% answered correctly on follow-up (**Table S1**).

In the sensitivity analysis, the fathers' scores still dropped significantly (as seen with the primary analysis) at the time of follow-up ($p<0.0001$). The mother's scores increased as noted in the initial analysis, and the difference now exceeded the threshold for significance ($p=0.0172$; original $p=0.0565$). The proband's scores increased, but the difference was still not statistically significant ($p=0.1175$). Overall, the entire group of respondents had a small but significant decrease in scores ($p=0.0027$) (**Table S1**).

Looking at the actual change in scores for the N=151 patients who completed the follow-up survey (**Table S2b**) the father's scores still decreased significantly ($p<0.0001$), the mothers' scores increased significantly ($p=0.0012$) and the probands scores did not change significantly ($p=0.500$). Overall for the entire cohort, there was a small decrease in scores and this was not statistically significant ($p=0.4196$).

### Table S1: Overall Comparison of Initial and Follow-up Scores[a], Excluding Patients Who Received Two Different Versions of Q1

| | Fathers | | | Mothers | | | Probands | | | Total | | |
|---|---|---|---|---|---|---|---|---|---|---|---|---|
| | Initial N=368 | Follow-up N=54 | pvalue[b] | Initial N=399 | Follow-up N=92 | pvalue[b] | Initial N=20 | Follow-up N=5 | pvalue[b] | Initial N=787 | Follow-up N=151 | pvalue[b] |
| Mean (SD) | 4.6 (0.8) | 3.6 (1.1) | <0.0001 | 3.7 (1.1) | 4.1 (0.8) | 0.0172 | 3.7 (1.2) | 4.6 (0.5) | 0.1175 | 4.1 (1.1) | 4.0 (1.0) | 0.0027 |
| Median | 5.0 | 4.0 | | 4.0 | 4.0 | | 3.5 | 5.0 | | 5.0 | 4.0 | |
| Q1, Q3 | 4.0, 5.0 | 3.0, 4.0 | | 3.0, 5.0 | 4.0, 5.0 | | 3.0, 5.0 | 4.0, 5.0 | | 4.0, 5.0 | 3.0, 5.0 | |
| Range | (1.0, 5.0) | (1.0, 5.0) | | (1.0, 5.0) | (2.0, 5.0) | | (2.0, 5.0) | (4.0, 5.0) | | (1.0, 5.0) | (1.0, 5.0) | |

[a] Score is the total number of correct answers out of 5 questions
[b] Wilcoxon rank sum test

### Table S2a: Overall Initial and Follow-up Question Responses, Excluding Patients Who Got Two Different Versions of Q1



| | | Fathers | | | Mothers | | | Probands | | | Total | | |
|---|---|---|---|---|---|---|---|---|---|---|---|---|---|
| Survey question | Correct Response | Initial N=152 | Follow-up N=29 | pvalue[a] | Initial N=163 | Follow-up N=52 | pvalue[a] | Initial N=9 | Follow-up N=4 | pvalue[a] | Initial N=324 | Follow-up N=85 | pvalue[a] |
| Do I have to be part of the GCCP study?[2] | No | 136 (89.5%) | 23 (79.3%) | 0.1248 | 91 (55.8%) | 47 (90.4%) | <0.0001 | 9 (100.0%) | 4 (100.0%) | . | 236 (72.8%) | 74 (87.1%) | 0.0064 |

[a] Wilcoxon rank sum test

| | | Fathers | | | Mothers | | | Probands | | | Total | | |
|---|---|---|---|---|---|---|---|---|---|---|---|---|---|
| Survey question | Correct Response | Initial N=216 | Follow-up N=25 | pvalue[a] | Initial N=236 | Follow-up N=40 | pvalue[a] | Initial N=11 | Follow-up N=1 | pvalue[a] | Initial N=463 | Follow-up N=66 | pvalue[a] |
| Is it my choice to be part of the GCCP study?[2] | Yes | 212 (98.1%) | 25 (100.0%) | 0.4926 | 234 (99.2%) | 40 (100.0%) | 0.5590 | 11 (100.0%) | 1 (100.0%) | . | 457 (98.7%) | 66 (100.0%) | 0.3523 |

[a] Wilcoxon rank sum test

### Table S2b: Comparison of Initial and Follow-up Scores[a] Among Respondents Who Completed Both Surveys, Excluding Patients Who Received Two Different Versions of Q1 (Score = number of correct answers out of 5 questions)

| | Fathers N=54 | | | | Mothers N=92 | | | | Probands N=5 | | | | Total N=151 | | | |
|---|---|---|---|---|---|---|---|---|---|---|---|---|---|---|---|---|
| | Initial | Follow-up | Change (Initial – Follow-up) | pvalue[b] | Initial | Follow-up | Change (Initial – Follow-up) | pvalue[b] | Initial | Follow-up | Change (Initial – Follow-up) | pvalue[b] | Initial | Follow-up | Change (Initial – Follow-up) | pvalue[b] |
| Mean (SD) | 4.7 (0.6) | 3.6 (1.1) | 1.0 (1.2) | <.0001 | 3.7 (1.1) | 4.1 (0.8) | -0.4 (1.2) | 0.0012 | 4.2 (1.1) | 4.6 (0.5) | -0.4 (1.5) | 0.5000 | 4.1 (1.1) | 4.0 (1.0) | 0.1 (1.4) | 0.4196 |
| Median | 5.0 | 4.0 | 1.0 | . | 4.0 | 4.0 | 0.0 | . | 5.0 | 5.0 | 0.0 | . | 4.0 | 4.0 | 0.0 | . |
| Q1, Q3 | 4.0, 5.0 | 3.0, 4.0 | 0.0, 2.0 | . | 3.0, 5.0 | 4.0, 5.0 | -1.0, 0.0 | . | 3.0, 5.0 | 4.0, 5.0 | -2.0, 1.0 | . | 3.0, 5.0 | 3.0, 5.0 | -1.0, 1.0 | . |
| Range | (3.0, 5.0) | (1.0, 5.0) | (-1.0, 4.0) | . | (1.0, 5.0) | (2.0, 5.0) | (-3.0, 2.0) | . | (3.0, 5.0) | (4.0, 5.0) | (-2.0, 1.0) | . | (1.0, 5.0) | (1.0, 5.0) | (-3.0, 4.0) | . |

[a] Score is the total number of correct answers out of 5 questions
[b] Wilcoxon rank sum test

**Supplemental Table 3.** Association Between Study Elements and Overall Satisfaction Scores[a] (Expanded Data)

| Factors | Overall satisfaction scores | | | | | Kruskal-Wallis P-value |
|---|---|---|---|---|---|---|
| | -2 Strongly dissatisfied | -1 Dissatisfied | 0 Neutral | 1 Satisfied | 2 Strongly satisfied | |
| **Comprehension[b]** | | | | | | |
| **1** | 0 (0.0%) | 0 (0.0%) | 0 (0.0%) | 1 (1.5%) | 1 (0.9%) | 0.0293 |
| **2** | 0 (0.0%) | 0 (0.0%) | 0 (0.0%) | 4 (6.1%) | 15 (13.4%) | |
| **3** | 1 (100.0%) | 0 (0.0%) | 0 (0.0%) | 5 (7.6%) | 14 (12.5%) | |
| **4** | 0 (0.0%) | 0 (0.0%) | 2 (50.0%) | 15 (22.7%) | 40 (35.7%) | |
| **5** | 0 (0.0%) | 2 (100.0%) | 2 (50.0%) | 41 (62.1%) | 42 (37.5%) | |
| **Retention[c]** | | | | | | |
| **1** | 0 (0.0%) | 0 (0.0%) | 0 (0.0%) | 0 (0.0%) | 2 (1.8%) | 0.2600 |
| **2** | 0 (0.0%) | 0 (0.0%) | 0 (0.0%) | 6 (9.1%) | 11 (9.8%) | |
| **3** | 0 (0.0%) | 0 (0.0%) | 0 (0.0%) | 13 (19.7%) | 20 (17.9%) | |
| **4** | 0 (0.0%) | 0 (0.0%) | 4 (100.0%) | 24 (36.4%) | 54 (48.2%) | |
| **5** | 1 (100.0%) | 2 (100.0%) | 0 (0.0%) | 23 (34.8%) | 25 (22.3%) | |
| **Clarity[d]** | | | | | | |
| **-2** | 0 (0.0%) | 0 (0.0%) | 0 (0.0%) | 1 (1.5%) | 0 (0.0%) | <.0001 |
| **-1** | 0 (0.0%) | 0 (0.0%) | 0 (0.0%) | 0 (0.0%) | 0 (0.0%) | |
| **0** | 0 (0.0%) | 1 (50.0%) | 2 (50.0%) | 5 (7.6%) | 0 (0.0%) | |
| **1** | 1 (100.0%) | 1 (50.0%) | 1 (25.0%) | 51 (77.3%) | 30 (26.8%) | |
| **2** | 0 (0.0%) | 0 (0.0%) | 1 (25.0%) | 9 (13.6%) | 82 (73.2%) | |
| **Adequacy of information[e]** | | | | | | |
| **-2** | 0 (0.0%) | 0 (0.0%) | 1 (25.0%) | 1 (1.5%) | 2 (1.8%) | 0.0552 |
| **-1** | 0 (0.0%) | 1 (50.0%) | 1 (25.0%) | 6 (9.1%) | 4 (3.6%) | |
| **0** | 1 (100.0%) | 1 (50.0%) | 2 (50.0%) | 59 (89.4%) | 106 (94.6%) | |
| **1** | 0 (0.0%) | 0 (0.0%) | 0 (0.0%) | 0 (0.0%) | 0 (0.0%) | |
| **2** | 0 (0.0%) | 0 (0.0%) | 0 (0.0%) | 0 (0.0%) | 0 (0.0%) | |
| **Privacy[f]** | | | | | | 0.0364 |
| **0** | 0 (0.0%) | 1 (50.0%) | 0 (0.0%) | 3 (4.5%) | 1 (0.9%) | |
| **1** | 1 (100.0%) | 1 (50.0%) | 4 (100.0%) | 63 (95.5%) | 111 (99.1%) | |

| Factors | Overall satisfaction scores | | | | | Kruskal-Wallis P-value |
|---|---|---|---|---|---|---|
| | -2 Strongly dissatisfied | -1 Dissatisfied | 0 Neutral | 1 Satisfied | 2 Strongly satisfied | |
| **Study goals**[g] | | | | | | |
| **-2** | 0 (0.0%) | 0 (0.0%) | 0 (0.0%) | 1 (1.5%) | 0 (0.0%) | <.0001 |
| **-1** | 0 (0.0%) | 0 (0.0%) | 0 (0.0%) | 0 (0.0%) | 0 (0.0%) | |
| **0** | 0 (0.0%) | 0 (0.0%) | 0 (0.0%) | 1 (1.5%) | 6 (5.4%) | |
| **1** | 0 (0.0%) | 2 (100.0%) | 4 (100.0%) | 42 (63.6%) | 34 (30.4%) | |
| **2** | 1 (100.0%) | 0 (0.0%) | 0 (0.0%) | 22 (33.3%) | 72 (64.3%) | |
| **Participant trust**[h] | | | | | | |
| **0** | 0 (0.0%) | 0 (0.0%) | 0 (0.0%) | 1 (1.5%) | 1 (0.9%) | 0.7122 |
| **1** | 0 (0.0%) | 0 (0.0%) | 0 (0.0%) | 6 (9.1%) | 10 (8.9%) | |
| **2** | 0 (0.0%) | 1 (50.0%) | 1 (25.0%) | 7 (10.6%) | 9 (8.0%) | |
| **3** | 0 (0.0%) | 1 (50.0%) | 1 (25.0%) | 21 (31.8%) | 31 (27.7%) | |
| **4** | 1 (100.0%) | 0 (0.0%) | 2 (50.0%) | 31 (47.0%) | 61 (54.5%) | |

[a] *Satisfaction* score was scored from strongly disagree (-2) to strongly agree (2) on the survey question “I had an easy time signing up for the Genetic Causes of CP study.”
[b] *Comprehension* score was defined as the number of comprehension questions answered correctly on the initial survey out of 5 possible.
[c] *Retention* score was defined as the number of comprehension questions answered correctly on the follow-up survey.
[d] *Adequacy of Information* was scored from not nearly enough (-2) to way too much (2) based on the statement “The amount of information I was given about the study was _________.”
[e] *Clarity* was scored from strongly disagree (-2) to strongly agree (2) on the survey question “When I joined the study, I understood what I had to do to sign up.”
[f] *Privacy* was scored as yes (1) or no (0) on the survey question “Are the privacy protections for the study strong enough?”
[g] *Study goal* was scored from strongly disagree (-2) to strongly agree (2) on the survey question “When I signed up for the study, I understood that the researchers are trying to find changes in my family's DNA that cause cerebral palsy.”
[h] *Participant trust* score was defined as the number of H-TBR scale questions answered as indicating trust. Answers indicating trust are as follows:

“Doctors who do medical research care only about what is best for each patient.” (Agree)
“Doctors tell their patients everything they need to know about being in a research study.” (Agree)
“Medical researchers treat people like "guinea pigs." (Disagree)
“I completely trust doctors who do medical research.” (Agree)
